# Aging and Alzheimer’s Disease Have Dissociable Effects on Medial Temporal Lobe Connectivity

**DOI:** 10.1101/2023.01.18.23284749

**Authors:** Stanislau Hrybouski, Sandhitsu R. Das, Long Xie, Laura E. M. Wisse, Melissa Kelley, Jacqueline Lane, Monica Sherin, Michael DiCalogero, Ilya Nasrallah, John A. Detre, Paul A. Yushkevich, David A. Wolk

## Abstract

Functional disruption of the medial temporal lobe-dependent networks is thought to underlie episodic memory deficits in aging and Alzheimer’s disease. Previous studies revealed that the anterior medial temporal lobe is more vulnerable to pathological and neurodegenerative processes in Alzheimer’s disease. In contrast, cognitive and structural imaging literature indicates posterior, as opposed to anterior, medial temporal lobe vulnerability in normal aging. However, the extent to which Alzheimer’s and aging-related pathological processes relate to functional disruption of the medial temporal lobe-dependent brain networks is poorly understood. To address this knowledge gap, we examined functional connectivity alterations in the medial temporal lobe and its immediate functional neighborhood – the Anterior-Temporal and Posterior-Medial brain networks – in normal agers, individuals with preclinical Alzheimer’s disease, and patients with Mild Cognitive Impairment or mild dementia due to Alzheimer’s disease. In the Anterior-Temporal network and in the perirhinal cortex, in particular, we observed an inverted ‘U-shaped’ relationship between functional connectivity and Alzheimer’s stage. According to our results, the preclinical phase of Alzheimer’s disease is characterized by increased functional connectivity between the perirhinal cortex and other regions of the medial temporal lobe, as well as between the anterior medial temporal lobe and its one-hop neighbors in the Anterior-Temporal system. This effect is no longer present in symptomatic Alzheimer’s disease. Instead, patients with symptomatic Alzheimer’s disease displayed reduced hippocampal connectivity within the medial temporal lobe as well as hypoconnectivity within the Posterior-Medial system. For normal aging, our results led to three main conclusions: (1) intra-network connectivity of both the Anterior-Temporal and Posterior-Medial networks declines with age; (2) the anterior and posterior segments of the medial temporal lobe become increasingly decoupled from each other with advancing age; and, (3) the posterior subregions of the medial temporal lobe, especially the parahippocampal cortex, are more vulnerable to age-associated loss of function than their anterior counterparts. Together, the current results highlight evolving medial temporal lobe dysfunction in Alzheimer’s disease and indicate different neurobiological mechanisms of the medial temporal lobe network disruption in aging vs. Alzheimer’s disease.

## INTRODUCTION

Structures of the Medial Temporal Lobe (MTL) have attracted extensive scientific inquiry for over fifty years because of this region’s role not only in episodic memory but also in the processing of emotional content, social cognition, creative thought, short-term memory, and object discrimination.^1–10^ Both human and animal studies implicate the MTL as the key region involved in memory decline in aging and Alzheimer’s disease (AD).^11–14^

Core memory-associated MTL subregions are the hippocampus (HP) and the perirhinal (PRC), entorhinal (ERC), and parahippocampal (PHC) cortices.^6,7,13^ Although unresolved, there is considerable evidence that the functional organization of the MTL can be divided along the anterior-posterior axis.^6,7,15^ Further, a growing body of clinical literature indicates the differential vulnerability of the anterior vs. posterior MTL to atrophy and pathological processes.^16–20^ The anterior MTL consists of the anterior HP (primarily head), PRC, and ERC, while the posterior MTL consists of the posterior (body + tail) HP and PHC. Because the MTL is one of the most densely connected regions in the brain,^21^ MTL dysfunction is likely to propagate to extra-MTL brain areas with which various MTL subregions interact. Thus, changes in MTL function ought to be considered in the context of their functional communities. Two brain networks have been hypothesized to explain MTL-cortical interactions: the Anterior-Temporal (AT) and Posterior-Medial (PM).^6,22–25^ The anterior MTL regions (i.e., PRC, ERC, anterior HP, and amygdala) belong to the AT network, which also includes a number of extra-MTL regions such as lateral temporal and orbitofrontal cortices, and the temporal pole.^6^ The PM network consists of the posterior MTL (i.e., posterior HP and PHC, but sometimes also includes medial ERC) along with a number of medial parietal regions, especially the posterior cingulate, precuneus, and retrosplenial.^6,26^ In AD, tau pathology is more pronounced in the AT network, while amyloid plaques are more prevalent in the PM network.^24,27^

MTL dysfunction is an early feature of AD,^11,12,28–31^ corroborated by previous task-based functional neuroimaging studies which reported increased HP activation in patients with Mild Cognitive Impairment (MCI),^32–36^ and in individuals at genetic risk for AD, including asymptomatic carriers of the apolipoprotein ε4 allele.^37–42^ Results from functional connectivity (FC) studies have been less clear with mixed findings of both increased and decreased MTL connectivity in patients with MCI or dementia-level impairment due to AD.^22,43–47^ In addition, changes in the MTL function at the systems level in normal aging are largely unexplored, and despite their relevance to declining memory in aging and AD, the network architecture of the AT and PM systems has not been precisely mapped. Given these knowledge gaps in the field, the primary aim of our current study was to determine whether and how age-associated FC changes within the MTL, and its immediate functional neighborhood – AT and PM network systems – differ from those that are caused by AD, especially during the earliest preclinical stage of the disease, when clinical interventions have the greatest potential. Our secondary goal was to construct detailed maps of MTL-associated networks.

As suggested by animal models of AD that revealed amyloid-induced hyperexcitability within the MTL circuitry, including subclinical seizures, during the early stages of the disease,^48–54^ we predicted MTL hyperconnectivity in individuals with preclinical AD. Since animal work has also demonstrated the activity-dependent nature of tau accumulation and spread,^49,55^ we hypothesized that the aforementioned hyperconnectivity effect would be most prominent in the anterior MTL – and the PRC in particular – because this region includes the transentorhinal cortex, the earliest locus of tau pathology in AD.^12,14,17,21,56^ Furthermore, because rising tau is associated with synapse loss, cell death, and circuit breakdown,^57–59^ we predicted that symptomatic stages of AD would be characterized by reduced, not excessive, FC levels. In normal aging, we expected to see a generalized loss of network integrity, consistent with previous observations in other systems,^60–66^ but perhaps more prominently in the PM network given more robust declines in cognitive function ascribed to this network in prior studies of cognitive aging.^6,7,67–71^

## MATERIALS AND METHODS

### Participants

In this cross-sectional study, we analyzed data from 179 individuals from the Aging Brain Cohort (ABC) study of the University of Pennsylvania Alzheimer’s Disease Research Center (Penn ADRC). These participants undergo annual cognitive evaluations, including psychometric testing as prescribed by the Uniform Data Set 3.0.^72^ Consensus diagnoses are reached by a team that includes neurologists and neuropsychologists using clinical history and cognitive scores. Amyloid status was determined by a visual read of amyloid PET scans (see below for acquisition parameters). An additional group of younger adults (< 60 years of age) were recruited beyond the ABC study to capture the full adult age span. Thus, four groups were formed: (1) 36 cognitively unimpaired (CU) participants < 60 years of age were classified as *normal young and middle-aged adults* [age range 23-59 years; mean Montreal Cognitive Assessment (MoCA)^73^ total score ± SD = 26.1 ± 3.7], (2) 80 Aβ-negative CU participants > 60 years of age were classified as *normal older adults* [age range 63-86 years; mean MoCA ± SD = 27.4 ± 2.0], (3) 23 A β-positive CU participants > 60 years of age were categorized as *preclinical AD* [age range = 65-87 years; mean MoCA ± SD = 26.9 ± 1.6], and (4) 40 Aβ-positive cognitively impaired (CI) participants were categorized as *symptomatic AD* [age range 57-87 years; mean MoCA ± SD = 20.3 ± 4.2]. In all analyses aimed at studying the effects of normal aging on MTL connectivity, the first two groups (i.e., normal young, middle-aged, and older adults) were merged into a single *normal agers* cohort. Additional information about our participants is presented in Table 1. The study was approved by the University of Pennsylvania Institutional Review Board.

**Table 1:** Demographic information for each group in the study. Amyloid-PET standard uptake value ratio (SUVR) is shown for the Florbetaben tracer only.

| | Young & Middle-Aged (*N* = 36) | Aβ- CU (*N* = 80) | Aβ+ CU (*N* = 23) | Aβ+ CI (*N* = 40) |
| --- | --- | --- | --- | --- |
| Age (years) |  |  |  |  |
| Mean ± SD | 39.2 ± 11.8 | 71.1 ± 4.9 | 75.2 ± 6.0 | 71.7 ± 6.9 |
| Range [min/max] | 23/59 | 63/86 | 65/87 | 57/87 |
| Race |  |  |  |  |
| (White/Black/Asian/Other) | 20/12/3/1 | 61/17/0/2 | 21/2/0/0 | 35/4/0/1 |
| Sex (males/females) | 14/22 | 29/51 | 7/16 | 23/17 |
| Education (years) ± SD | 16.2 ± 2.6 | 16.6 ± 2.6 | 16.9 ± 1.7 | 16.5 ± 2.6 |
| Total MoCA ± SD | 26.1 ± 3.7 | 27.4 ± 2.0 | 26.9 ± 1.6 | 20.2 ± 4.3 |
| CDR (sum/global) | N/A | 0.038/0.031 | 0.087/0.065 | 2.962/0.638 |
| Composite Aβ SUVR ± SD (Florbetaben *n*) | N/A | 0.950 ± 0.062 (*n* = 68) | 1.280 ± 0.201 (*n* = 21) | 1.438 ± 0.144 (*n* = 37) |

### Image Acquisition

All structural and functional MRI datasets were acquired using a 3.0 T Siemens Prisma scanner (Erlangen, Germany) at the Center for Advanced Magnetic Resonance Imaging and Spectroscopy (University of Pennsylvania, Philadelphia, PA). During a resting-state functional MRI (rs-fMRI) scan, 420 functional volumes were collected axially using a T2*-sensitive Gradient Echo Planar Imaging (EPI) pulse sequence sensitive to blood oxygenation level-dependent (BOLD) contrast [repetition time (TR): 720 ms; echo time (TE): 37 ms; flip angle: 52°; field of view (FOV): 208×208 mm^2^; voxel size: 2×2×2 mm^3^; 72 interleaved slices; phase encoding direction: anterior to posterior; partial *k*-space: 7/8; multi-band acceleration factor: 8]. A whole-brain T1-weighted 3D Magnetization Prepared Rapid Gradient Echo (MPRAGE) sequence [TR: 2400 ms; TE: 2.24 ms; inversion time: 1060 ms; flip angle: 8°; FOV: 256×240×167 mm^3^; voxel size: 0.8×0.8×0.8 mm^3^] was used to acquire anatomical images for MTL subregion segmentation and registration to template space. To estimate B_0_ inhomogeneity, gradient echo field maps were also collected [TR: 580 ms; TE1/TE2: 4.12/6.58 ms; flip angle: 45°; FOV: 240×240 mm^2^; voxel size: 3×3×3 mm^3^; 60 interleaved slices].

Furthermore, all CU participants who were 60 years of age or older and all patients with symptomatic MCI or early AD underwent amyloid PET scanning. Our amyloid PET protocol used an injection of 8.1 mCi ± 20% of ^18^F-Florbetaben or 10 mCi ± 20% for ^18^F-Florbetapir and a 20 min brain scan (4 frames of 5-minute duration) following a standard uptake phase (a 90-min for ^18^F-Florbetaben and 50 min for ^18^F-Florbetapir). 44% of PET scans were acquired within six months of fMRI and 85% within a year of fMRI. Cognitively normal young and middle-aged adults underwent rs-fMRI but not PET imaging because this age group is unlikely to have measurable amyloid pathology.

### Image Preprocessing

Initial processing of structural and functional images was performed on the Flywheel Imaging Informatics Platform (http://flywheel.io). We used a customized *fMRIprep* 20.0.7^74^ analysis gear, which consisted of *mcflirt* realignment,^75,76^ susceptibility distortion correction using a custom workflow of *SDCFlows*,^77^ functional-to-structural boundary-based rigid-body registration,^78^ SyN diffeomorphic registration to the 2-mm MNI152 template using Advanced Normalization Tools (ANTs),^79,80^ followed by volumetric smoothing with a 6-mm FWHM Gaussian kernel, and decomposition of the fMRI time series into signal sources using the Probabilistic Spatial Independent Component Analysis (ICA).^81^

Next, manual ICA-based denoising was performed by a single rater (SH) with the aid of the automated movement component classifier, ICA-AROMA.^82^ Protocols and reliability of manual ICA-based artifact detection have been detailed elsewhere.^64,83,84^ Following the ICA-based denoising, the first 10 time points were discarded, and the rest of the fMRI pre-processing was done in the Conn toolbox (v. 20.b)^85^ for MATLAB (release 2020b; The MathWorks Inc., Natick, MA). Rather than employing global signal regression, which can introduce artifactual anti-correlations into the fMRI time series,^86–88^ we used 32 CompCorr^89^ regressors – 16 from the White Matter (WM) and 16 from the cerebrospinal fluid (CSF) – that were extracted from the partially de-noised fMRI data using eroded tissue probability maps. Those 32 CompCorr regressors were supplemented with movement regressors based on the Friston 24-Parameter model^90^ as well as low-frequency sine and cosine waves (6 waveforms with periods of π, 2π, and 3π over 410 fMRI volumes) that were included to account for slow BOLD signal drifts. Since realignment parameters in sub-second fMRI acquisitions are contaminated by breathing-associated magnetic field perturbations,^91,92^ those parameters were filtered using MATLAB scripts provided by Gratton *et al*.^91^ prior to being used as nuisance covariates. In the last step of our pre-processing pipeline, the fMRI time series were band-pass filtered (0.008-0.150 Hz) and underwent linear detrending. Descriptions of our *fMRIprep* structural pipeline as well as our amyloid-PET preprocessing pipeline are provided in the Supplementary Materials.

The MTL subregions were segmented using the automated segmentation of hippocampal subfields-T1 (ASHS-T1) pipeline.^93^ We used ASHS-T1 to generate Brodmann Area 36 (BA36), Brodmann Area 35 (BA35), ERC, PHC, as well as the anterior and posterior HP (aHP and pHP, respectively) segmentations. Because the theoretical framework tends to treat the PRC as a single functional unit,^6^ we merged the BA35 and BA36 ROIs into a single PRC ROI. For visual reference, a single participant’s ASHS-T1 segmentation is shown in Fig. 1a. Each subject’s ASHS-T1 MTL segmentations were then registered to MNI space using the same deformation fields that were used to register that subject’s fMRI data to the 2-mm MNI152 template. The warped ASHS-T1 ROIs were used to extract the denoised fMRI time series from each memory-related region of the MTL.

**Figure 1.**
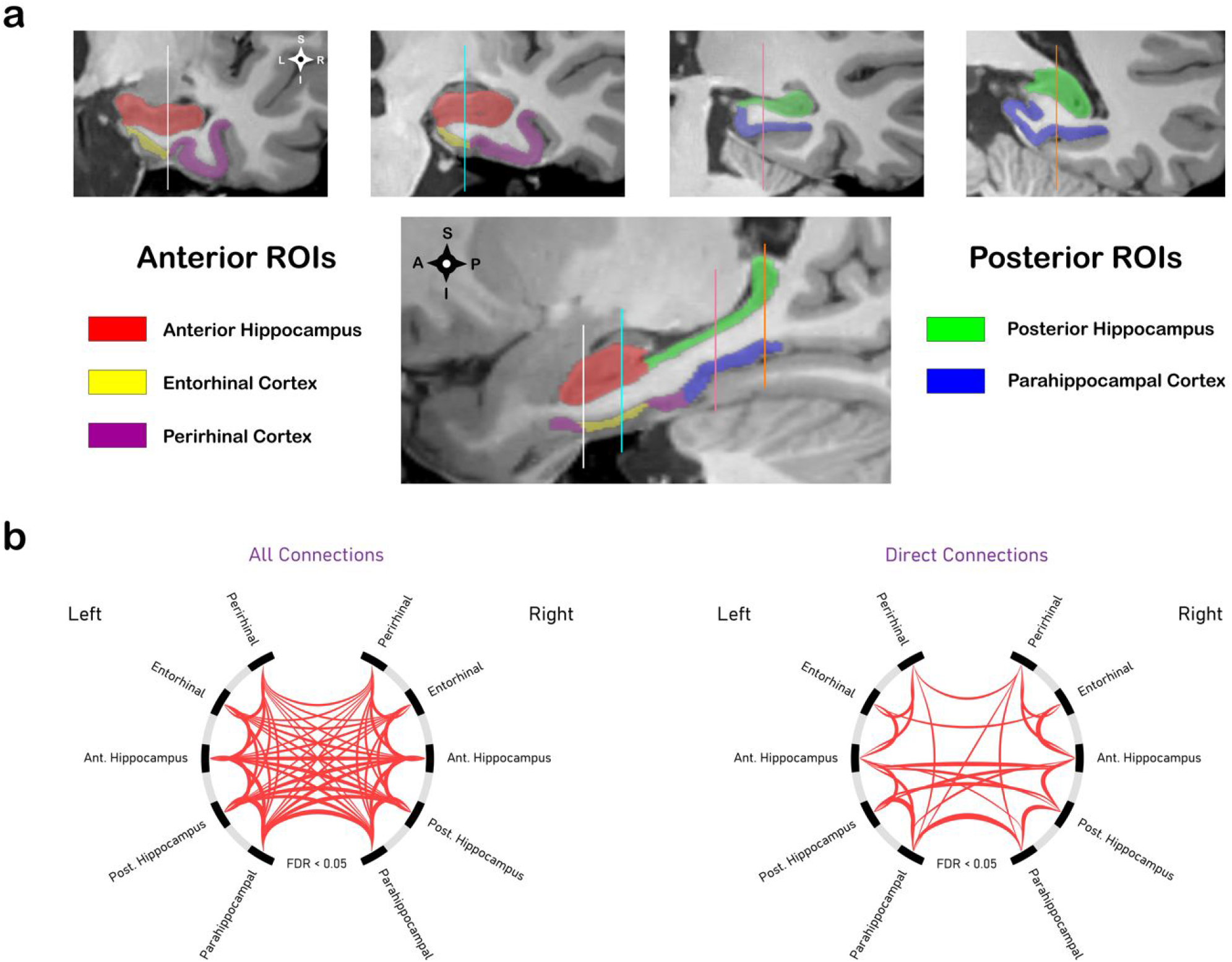
MTL subregions and their functional interactions with each other. **(a)** An example of MTL segmentation using ASHS-T1 pipeline; **(b)** Intra-MTL functional connectivity profiles in Aβ-negative cognitively normal adults. The connectogram on the left is based on correlational connectivity matrices, which do not control for indirect paths. The connectogram on the right is based on partial correlation matrices and represents direct functional interactions amongst various MTL subregions. Edge thickness in both connectograms is proportional to *t*-scores. Abbreviations: MTL = medial temporal lobe; FDR = false discovery rate.

### Intra-MTL Functional Connectivity Analyses

The most common technique for modeling the brain’s functional architecture is to use the bivariate Pearson correlation coefficient as a proxy for FC between brain regions.^94,95^ Despite its high sensitivity, this approach may produce biased connectivity estimates because it does not control for indirect paths.^95,96^ Such problems can be ameliorated by replacing bivariate correlation coefficients with partial correlations, controlling for signals from all other ROIs in the network model. However, the improvement in architectural accuracy comes at a price: noisier connectivity estimates and reduced statistical power when studying group effects.^96^ Because each FC metric has a unique set of trade-offs, we examined the effects of age and AD pathology on intra-MTL FC using both approaches.

All statistical analyses of intra-MTL FC were performed using the Network-Based Statistics (NBS) toolbox.^97^ The following NBS settings were used: (1) uncorrected connection-level *t*-statistic threshold was set to match two-tailed *p* < 0.05; (2) the cluster-mass ‘intensity’ option in the NBS algorithm was turned on; (3) positive and negative contrasts were run separately, and family-wise error (FWE)-corrected significance threshold was set at *p* < 0.025 for each one-sided (i.e., positive or negative) test; (4) 25,000 Freedman and Lane^98^ permutations were used to construct each cluster-mass distribution under the null hypothesis. Prior to performing age effect analyses and group comparisons of intra-MTL FC, correlation matrices were Fisher Z-transformed, and overall intra-MTL connectivity maps were generated using all participants from the *normal agers* cohort [positive-sided one-sample connection-level *t*-tests with the False Discovery Rate (FDR)^99^ correction for multiple comparisons]. These overall connectivity maps were used to select edges of interest (Fig. 1b). Since all MTL ROIs were functionally related to each other in the bivariate correlational method, all pairwise connections between ASHS-T1 ROIs were examined for the effects of age and AD pathology (Fig. 1b, left panel). For partial correlations, the above test identified 23 (out of 45) edges representing direct functional interactions within the MTL (Fig. 1b, right panel). In all subsequent intra-MTL FC analyses that used partial correlations, only those 23 edges were studied.

To study the effects of age on intra-MTL FC, we ran a General Linear Model (GLM) on FC data from the *normal agers* cohort with sex and 4 aggregate head movement metrics [mean filtered framewise displacement (FD), maximum filtered FD, mean DVARS, max DVARS; see Power *et al*.^100^ for metric details] as nuisance covariates. To examine how MTL function differs between normal agers and individuals at different stages of AD progression, we performed all pairwise group comparisons of intra-MTL connectivity between *normal older adults, preclinical AD*, and *symptomatic AD*. In these group comparisons, age, sex, and aggregate head movement metrics were used as covariates. Connection-specific and supplementary statistical comparisons were performed using SPSS v. 28 (IBM Inc., Armonk, NY).

### Identification of Cortical Regions with Functional Connectivity to the Medial Temporal Lobe

Seminal work by Braak and Braak^17^ demonstrated that tau neurofibrillary tangle (NFT) pathology originates at the border of the PRC and ERC in the anterior MTL, and a recent *ex vivo* study by our group revealed the presence of an anterior-posterior NFT gradient within the MTL with posterior subregions being less vulnerable to NFT accumulation.^20^ Regions with higher/lower NFT burden scores had a remarkable degree of spatial overlap with the MTL portion of the AT/PM network, and together with the AT network’s vulnerability to tauopathy vs. PM network’s vulnerability to amyloidosis, seem to reflect partially dissociable patterns of molecular pathology within the anterior vs. posterior MTL and might also relate to dissociable patterns of network disruption in the AT vs. PM system.^6,22,24,27^ Thus, when modeling MTL interactions with the rest of the cortex, we employed four tau-based MTL ROIs (left/right × anterior/posterior). Our group recently built a novel atlas of MTL NFT accumulation using fusion of *ex vivo* MRI and serial histological imaging (Suppl. Fig. 1).^20^ Left and right tau-based MTL ROIs were defined by thresholding and binarizing this NFT accumulation map (inclusion threshold: WildCat-based metric of NFT burden > 0.2; for threshold details see Yushkevich *et al*.),^20^ and the anterior/posterior split was defined using the aHP/pHP boundary of the ASHS-T1 protocol.^7,93,101^ Next, we identified cortical regions with positive FC to at least one of the four tau-based MTL ROIs in the *normal agers* cohort (Fig. 2a). In ROI form, this extended MTL network was represented by the anterior and posterior tau-based MTL ROIs (aMTL_tau_ and pMTL_tau_, respectively) and 221 ROIs from the 400-region 17-Network Schaefer *et al*.^102^ parcellation (Fig. 2a; Suppl. Materials).

**Figure 2.**
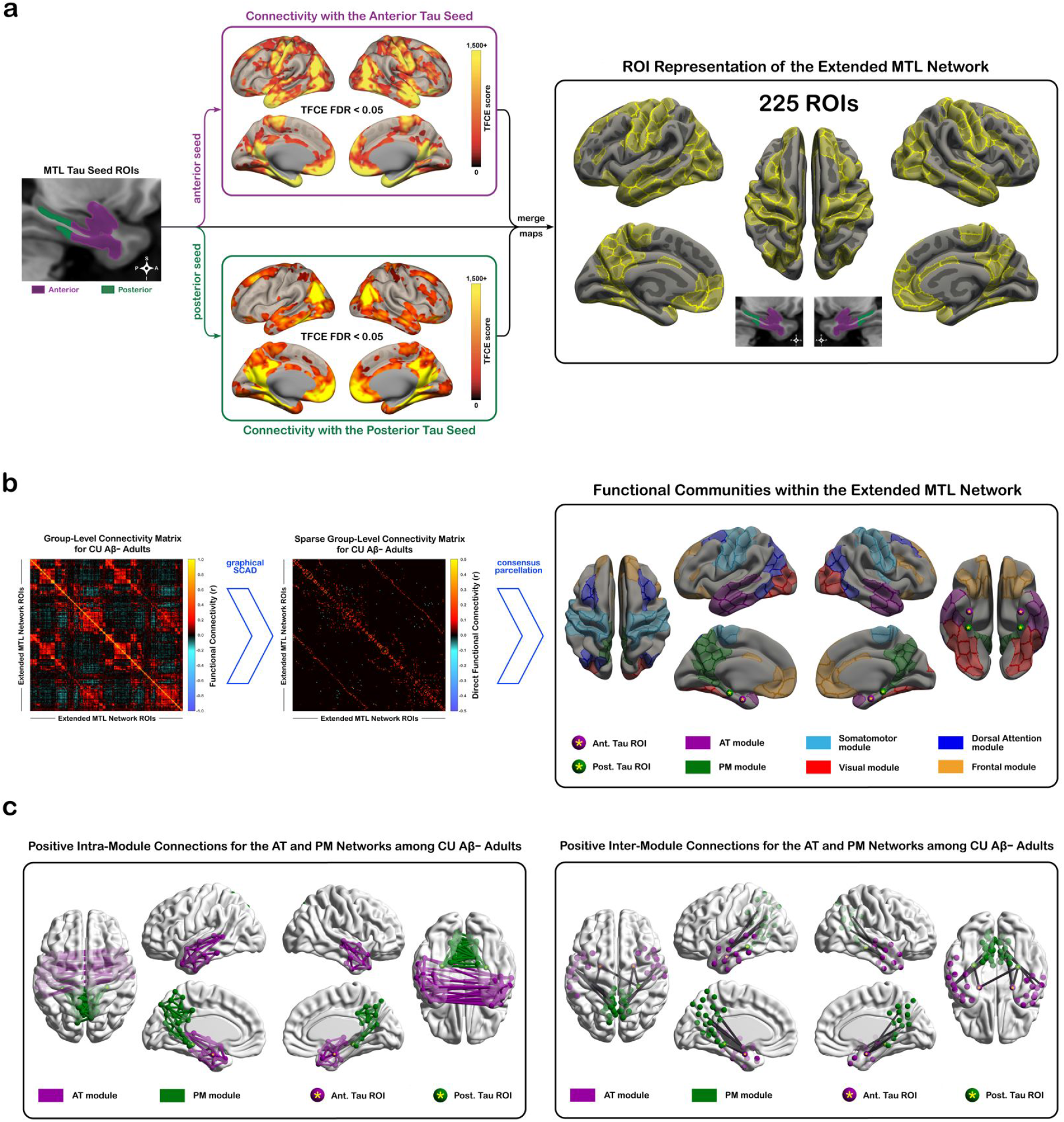
AT and PM network estimation. **(a)** Simplified schematic of the methodology that was used to select cortical regions that constitute the extended MTL network. Cortical regions with positive functional connectivity to the anterior or posterior tau-based MTL ROIs were identified and mapped onto ROI-based representation. **(b)** MTL network structure was estimated using graphical SCAD with BIC-based model selection. Functional modules within the extended MTL network were identified using consensus Louvain modularity. **(c)** AT and PM network architecture in the *normal agers* group (i.e., CU young, middle-aged, and Aβ-negative older participants). Left panel depicts the intra-module connections of each network; right panel depicts AT-PM inter-module connections. Abbreviations: AT = anterior-temporal; PM = posterior-medial; MTL = medial temporal lobe; CU = cognitively unimpaired; Ant. = anterior; Post. = posterior; TFCE = threshold-free cluster enhancement; FDR = false discovery rate; SCAD = smoothly clipped absolute deviation estimator.

### Estimating MTL Network Architecture

The functional architecture of the extended MTL network was estimated using the graphical SCAD algorithm.^103–106^ Similar to other sparse estimation techniques, graphical SCAD eliminates spurious or indirect connections by penalizing excessive model complexity if there is a lack of evidence in the data to support a more complex graph. Because sparse network estimation based on 225 ROIs and a relatively short fMRI acquisition is unlikely to produce sufficiently stable connectome estimates at the subject level, we performed group-level sparse network estimation instead. First, bivariate correlation matrices were computed for all *normal agers*.

Those correlation matrices were then Fisher-transformed and averaged such that each decade of human lifespan had equal weight on the final connectivity structure. The resulting average Z-connectivity matrix was converted into the correlational connectivity matrix and used as the covariance source in SCAD-based network estimation (Fig. 2b). Graphical SCAD relies on two tuning parameters: *α* and *ρ*. To minimize the Bayes risk, Fan and Li^104^ recommend *α* = 3.7. The second tuning parameter, *ρ*, was selected from a set of *ρ* = {*e*^−8.0^, *e*^−7.8^, *e*^−7.6^, …, *e*^0^} by minimizing the Bayesian Information Criterion (BIC).^105,106^ Custom MATLAB scripts, employing the QUIC optimizer,^107^ were used to solve the graphical SCAD problem (Fig. 2b; Suppl. Materials).

To identify the AT and PM network communities within the broader MTL connectome, we used the two-sided Louvain modularity algorithm that incorporates both positive and negative edge weights in its community search.^108,109^ Since modularity-based network parcellations can produce inconsistent networks from one iteration to another, we built a consensus parcellation from 5,000 separate iterations of the fine-tuned two-sided Louvain community search.^109–111^ Module detection was performed on the *normal agers’* sparse partial correlation matrix estimated above. In total, we identified 6 stable functional modules that could explain functional relationships among the 225 MTL-affiliated ROIs (Fig. 2b). Two of those modules were consistent with the AT and PM networks that have been reported in the literature.^6^ The parcellation procedure was performed using functions from the Brain Connectivity Toolbox.^112^

### Statistical Analyses of Intra-AT, Intra-PM, and AT-PM Connectivity

Age effect analyses and AD group comparisons were performed on bivariate connectivity matrices, but only on those connections that survived group-level graphical SCAD estimation in at least one test-related group. For age effect analyses, in addition to the overall connectivity map that was estimated above, we also estimated separate SCAD-based maps for younger (*normal agers* < 60 years) and older (*normal agers* > 60 years) participants. If a connection was positive in at least one of the three (i.e., younger, older, overall) group-level sparse connectivity matrices, it was examined for age effects. Negative edges and those with conflicting signs in the bivariate vs. graphical SCAD models were excluded because of interpretability issues. The final set of intra-AT, intra-PM, and AT-PM connections that were used in age effect analyses is shown in Fig. 2c. For AD group comparisons, similar criteria were used, except that there were 2 instead of 3 group-level connectivity estimates per comparison. For example, for the preclinical vs. symptomatic AD connectivity comparison, two separate group-level graphical SCAD connectivity estimates were computed: the first for the preclinical group and the second for the symptomatic group.

Statistical analyses of the AT and PM intra-module and AT-PM connectivity were performed using the NBS GLM approach^97^ with all settings, including covariates, matching those that were used in the intra-MTL analyses described above. Connection-specific follow-up comparisons were performed using SPSS v. 28 (IBM Inc., Armonk, NY). All AT and PM network results were visualized with the BrainNet Viewer.^113^

### Data Availability

Anonymized preprocessed data and in-house analysis scripts will be made available upon request for the sole purpose of replicating procedures and results presented in this article.

## RESULTS

### Effects of Age and AD Progression on Functional Connectivity between the MTL Subregions

The NBS algorithm identified a single cluster of 14 edges that represents declining FC in *normal agers* (FWE *p* < 0.01; Fig. 3a). Most of the cluster’s connections were interhemispheric (10 out of 14 edges), and all but one represented either anterior-with-posterior or posterior-with-posterior functional interactions. The PHC was involved in 9 out of 14 connections with a negative correlation to age. We did not observe any positive associations between age and intra-MTL FC.

**Figure 3.**
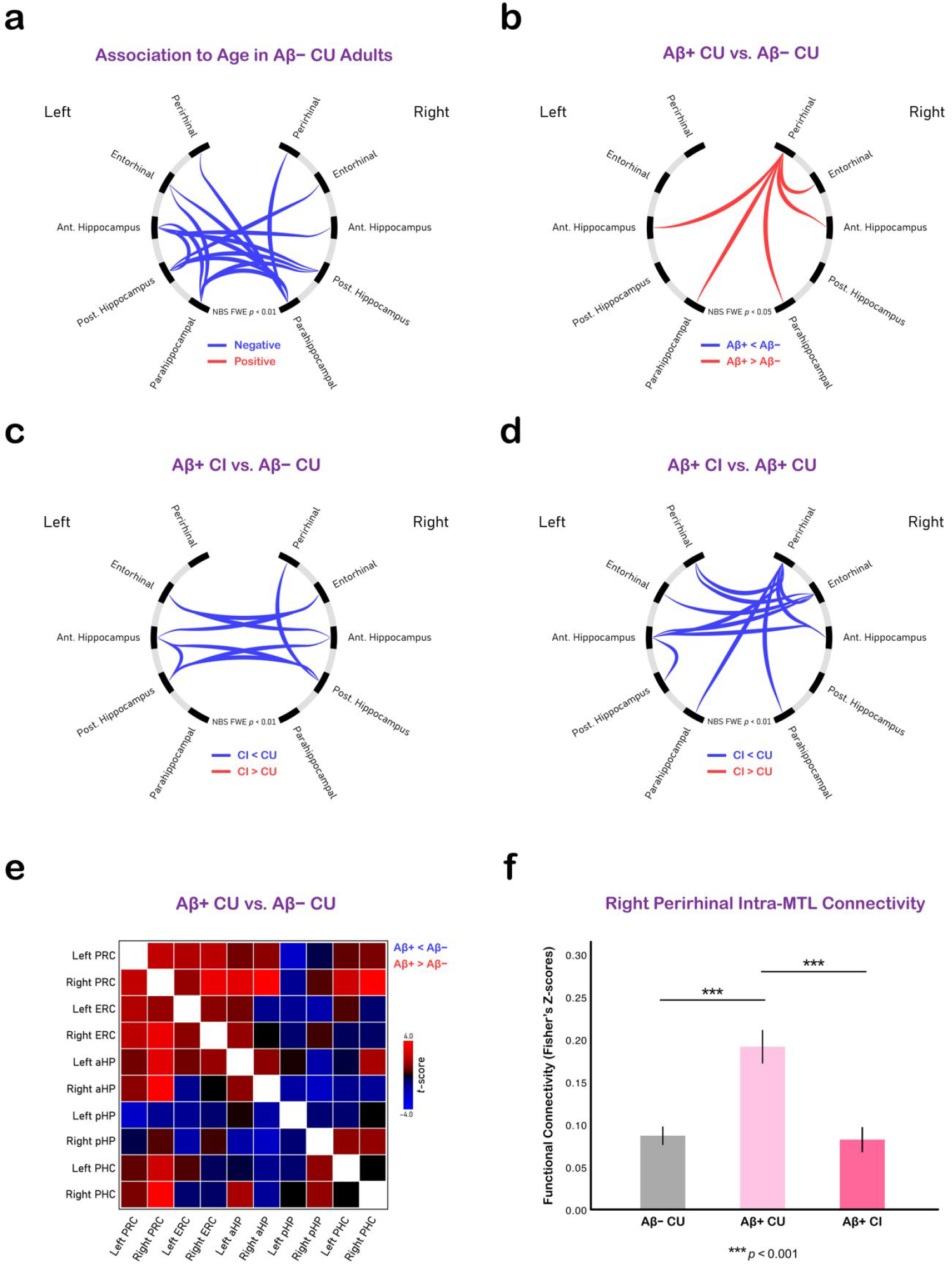
Connectograms representing the effects of **(a)** age and **(b-d)** AD progression on intra-MTL functional connectivity. **(e)** Matrix-form representation of intra-MTL functional connectivity differences between Aβ-positive individuals with preclinical AD and Aβ-negative age-matched controls. **(f)** Right PRC functional connectivity differences between age-matched Aβ-negative normal agers, Aβ-positive cognitively normal individuals with preclinical AD, and Aβ-positive individuals with symptomatic disease. Abbreviations: PRC = perirhinal cortex; ERC = entorhinal cortex; PHC = parahippocampal cortex; aHP = anterior hippocampus; pHP = posterior hippocampus; CU = cognitively unimpaired; CI, cognitively impaired; NBS = network-based statistic.

Comparing intra-MTL connectivity profiles of the Aβ− CU and Aβ+ CU older adult groups revealed increased functional coupling between the right PRC and other MTL subregions in the preclinical AD group (Fig. 3b). Although PRC hyperconnectivity reached statistical significance in the right hemisphere only, similar trends were observed in the left hemisphere as well (Fig. 3e). To test whether these alterations in MTL function were linked to structural atrophy in the MTL, we used two-way ANCOVAs [Group and Sex as fixed factors; Age as a covariate] to compare the Aβ− CU vs. Aβ+ CU groups MTL volumetric measurements (thickness for PRC, ERC, and PHC; ICV-corrected volumes for aHP and pHP; thickness/volumes averaged across hemispheres). Because we did not detect any significant differences in MTL structure when comparing normal older adults to individuals with preclinical AD (all *p* values > 0.10), we concluded that MTL network dysfunction likely precedes neuronal loss in AD.

To investigate the effects of disease progression on the MTL function, we compared intra-MTL connectivity profiles of the Aβ+ CI group to those of the age-matched Aβ− CU and Aβ+ CU groups. Relative to normal older adults, individuals in the Aβ+ CI group displayed lower hippocampal connectivity (Fig. 3c), suggesting that functional changes in the symptomatic disease are characterized by functional decoupling of the two hippocampi from each other and from the rest of the MTL. Contrasting intra-MTL connectivity profiles of the Aβ+ CI group with those of the preclinical AD group also revealed a cluster of hypoconnectivity in the symptomatic group. This cluster consisted of 11 connections, 9 of which represented either PRC-or ERC-associated connectivity (Fig. 3d).

Taken together, the above results suggest that the PRC (and possibly ERC) connectivity follows an inverse ‘U-shaped’ pattern that is characterized by an initial rise during the preclinical phase and a decline during the symptomatic stage. To verify this effect, we computed mean connectivity scores for the right PRC connections that displayed hyperconnectivity in the preclinical vs. control comparison above and examined how those PRC connectivity profiles differed at various stages of the AD continuum. Group comparisons [two-way ANCOVAs; Group and Sex as fixed factors; Age and Head Movement metrics as covariates] of these overall PRC connectivity measures showed that intra-MTL PRC connectivity was higher in the Aβ+ CU group than in the other two groups [Aβ+ CU vs. Aβ− CU comparison: *F*_1,91_ = 25.90, Holm-Bonferroni-corrected *p* < 0.001; Aβ+ CI vs. Aβ+ CU comparison: *F*_1,54_ = 25.89, Holm-Bonferroni-corrected *p* < 0.001]. However, right PRC connectivity of the Aβ− CU group did not differ from that of the Aβ+ CI group (uncorrected *p* = 0.95). Thus, our results indicate excess functional synchronicity in the anterior MTL only during the asymptomatic stage of the disease [age-, gender-, and motion-corrected PRC connectivity means in Fisher’s *Z*-score units: *M*_Aβ− CU_ = 0.085, *M*_Aβ+ CU_ = 0.209, *M*_Aβ+ CI_ = 0.085; Fig. 3f].

In summary, our intra-MTL connectivity comparisons revealed PRC hyperconnectivity in cognitively normal amyloid-positive individuals and HP hypoconnectivity in patients with symptomatic disease. In normal agers, we observed age-associated FC decline that was centered around the PHC connections. Furthermore, these trends did not change if partial correlations were used to model intra-MTL functional interactions (Suppl. Fig. 2).

### The Effects of Age and AD Progression on the AT and PM Networks

Both network systems displayed declining intra-network FC as a function of age, while no positive relationships to age were detected in either system (Fig. 4a). Relationships to age within the AT and PM systems were represented by single 20-edge and 40-edge clusters, respectively. Only the AT network’s cluster contained an edge with a direct link to one of the tau-based MTL ROIs (Fig. 4a). In analyses of AT-PM inter-network FC only the aMTL_tau_-pMTL_tau_ edge in the left hemisphere had a statistically significant negative relationship to age (Suppl. Fig. 3), demonstrating that FC between the anterior and posterior MTL segments declines with age. We did not observe any statistically significant positive relationships between age and AT-PM inter-module FC.

**Figure 4.**
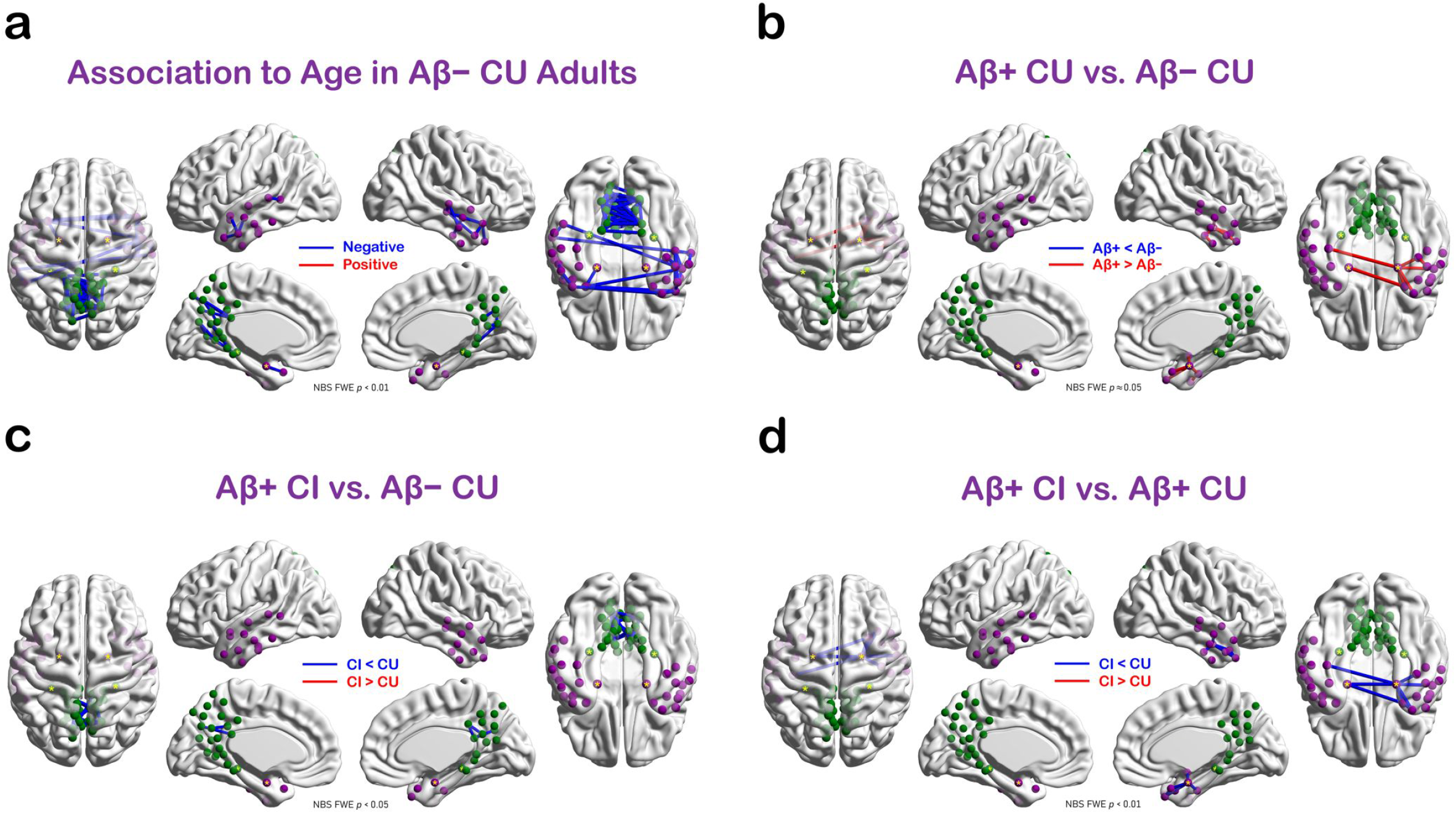
Effects of **(a)** age and **(b-d)** AD progression on intra-network connectivity of the AT and PM systems. AT nodes are in purple; PM nodes are in green. Tau-based MTL ROIs are marked with a yellow asterisk. Abbreviations: AT = anterior-temporal; PM = posterior-medial; CU = cognitively unimpaired; CI, cognitively impaired.

Consistent with the intra-MTL effects detailed above, the preclinical AD group displayed AT-specific hyperconnectivity relative to age-matched amyloid-negative controls. This excess connectivity was represented by a single cluster of connections, all but one of which emanated from the aMTL_tau_ ROIs. As in the intra-MTL results, the AT-specific hyperconnectivity was more prominent in the right hemisphere and was not present in the symptomatic group (Fig. 4b). No FC differences between Aβ− CU and the Aβ+ CU groups were detected in the PM system (Fig. 4b). Relative to normal agers, patients with MCI and prodromal AD displayed reduced FC within the PM, but not the AT, system (Fig. 4c). Furthermore, in congruence with our intra-MTL findings, our analyses of the AT and PM networks indicate that hyperconnectivity within the AT system declines as pathology and symptoms progress beyond the preclinical stage (Fig. 4d). No differences in the AT-PM inter-network FC were found between the Aβ− CU, Aβ+ CU, and Aβ+ CI groups.

In order to further examine the effects of age on those connections that link the aMTL_tau_ and pMTL_tau_ ROIs to their respective networks, we first computed average functional connectivity scores of all such connections (i.e., one-hop aMTL_tau_-AT and pMTL_tau_-PM connectivity; Figs. 5–6). In normal agers, both the aMTL_tau_-AT and pMTL_tau_-PM one-hop FC was negatively associated with age (Fig. 5). Next, we evaluated the effect of disease progression on the aMTL_tau_-AT and pMTL_tau_-PM connectivity (Fig. 6). Disease stage group differences were statistically significant for the aMTL_tau_-AT connections (*F*_2,131_ = 9.17, *p* < 0.001), and showed a trend towards statistical significance for the pMTL_tau_-PM connections (*F*_2,131_ = 2.68, *p* = 0.072). Follow-up pairwise comparisons revealed that the aMTL_tau_-AT FC was greater in the Aβ+ CU group relative to the A β− CU control group [*F*_1,131_ = 11.98, Holm-Bonferroni-corrected FWE *p* = 0.001, *M*_diff_ = 0.084 Fisher’s *Z*-score units; Fig. 6] and relative to the Aβ+ CI patient group [*F*_1,131_ = 17.76; Holm-Bonferroni-corrected *p* < 0.001, *M*_diff_ = 0.114 Fisher’s *Z*-score units; Fig. 6]. No statistical differences in the aMTL_tau_-AT connectivity were observed between the Aβ− CU and Aβ+ CI groups (uncorrected *p* = 0.148; Fig. 6). For the pMTL_tau_-PM analyses, we did not find an increase in Aβ+ CI connectivity relative to the other groups. However, we observed a trend towards lower connectivity in the Aβ+ CI group relative to the Aβ− CU group [*F*_1,131_ = 5.16, Holm-Bonferroni-corrected FWE *p* = 0.074; Fig. 6].

**Figure 5.**
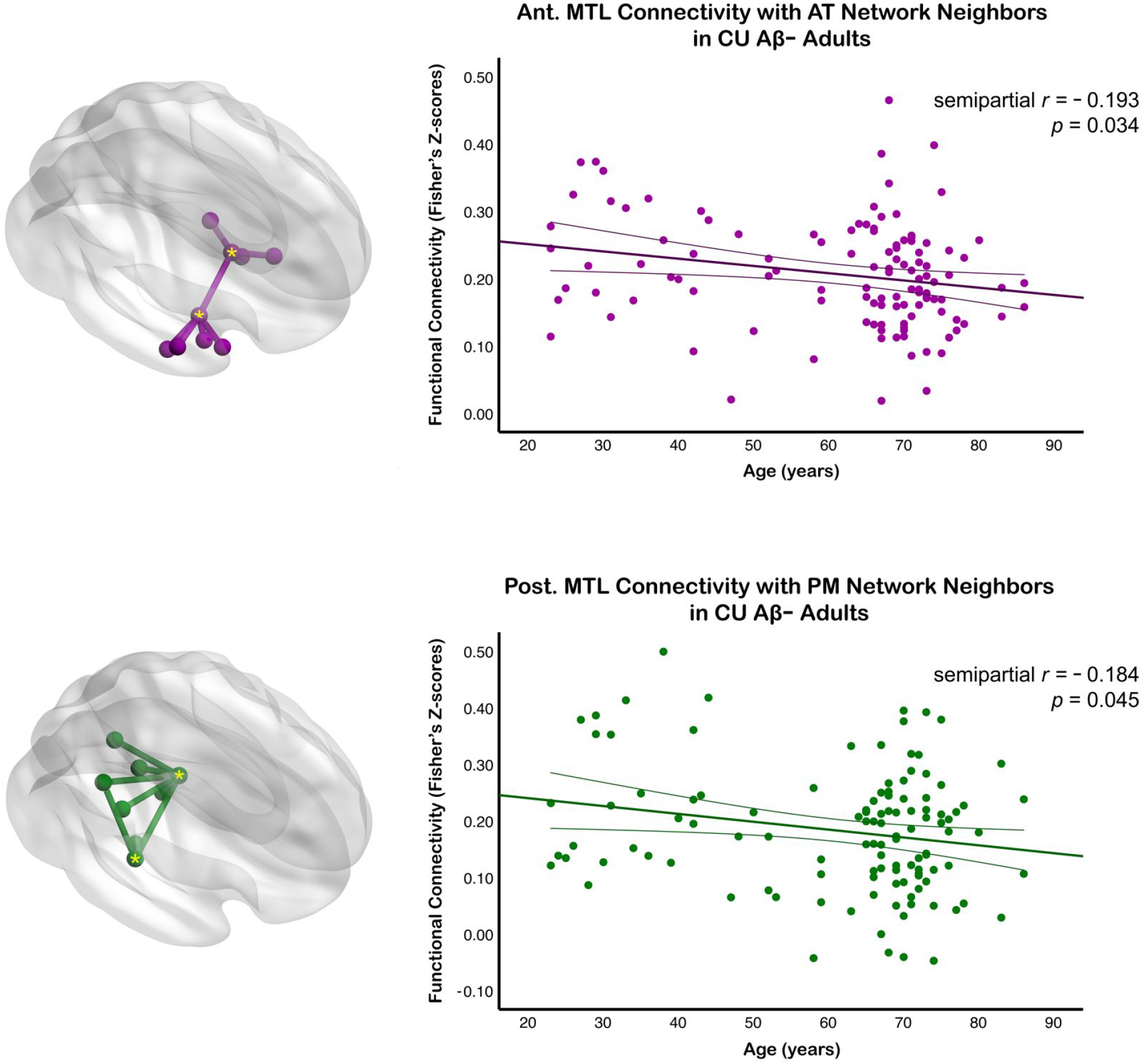
Age relationships for aMTL_tau_-AT and pMTL_tau_-PM functional connectivity. The aMTL_tau_-AT/pMTL_tau_-PM connectivity metric was computed by averaging connectivity values of those edges that represented direct functional interactions between the aMTL_tau_ (or pMTL_tau_) ROIs and their one-hop neighbors in the AT (or PM) network (top and bottom glass brains, respectively). Lifespan trajectories are surrounded by thin lines that represent 95% confidence intervals. The AT network’s nodes and edges are shown in purple; those of the PM network are shown in green. Tau-based MTL ROIs are marked with a yellow asterisk. Abbreviations: AT = anterior-temporal; PM = posterior-medial; MTL = medial temporal lobe; aMTL_tau_ = tau-based anterior MTL ROI; pMTL_tau_ = tau-based posterior MTL ROI; CU = cognitively unimpaired; Ant. = anterior; Post. = posterior.

**Figure 6.**
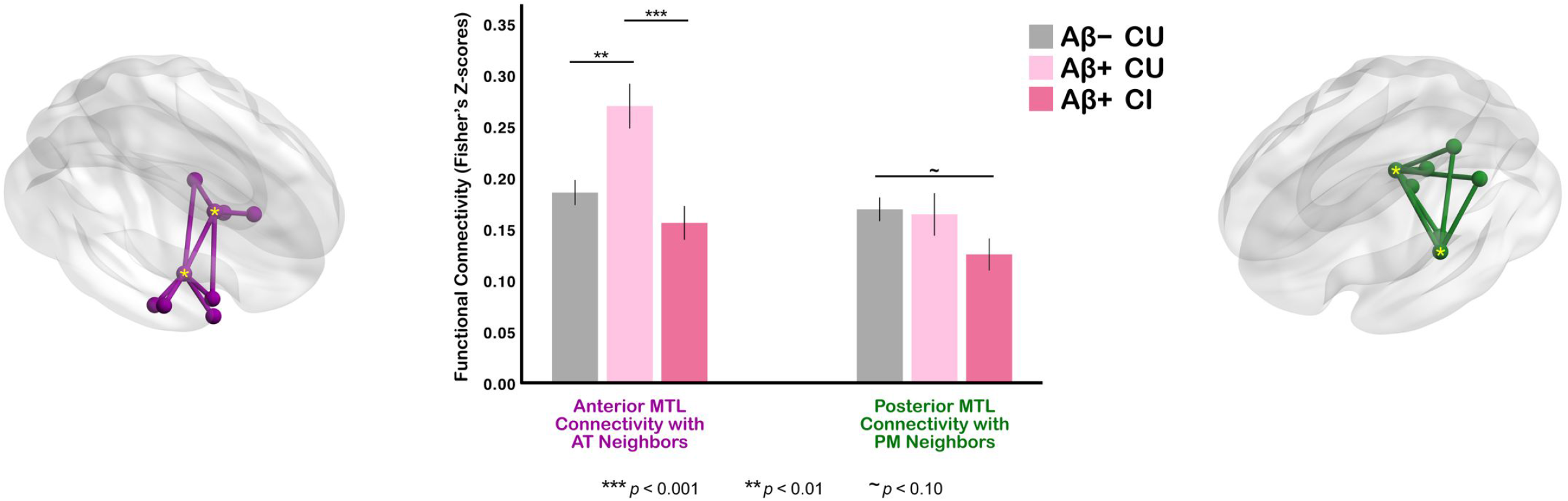
Effect of AD progression on aMTL_tau_-AT and pMTL_tau_-PM functional connectivity. The bar plot depicts statistical differences in the aMTL_tau_-AT and pMTL_tau_-PM connectivity between Aβ-negative cognitively unimpaired older adults, Aβ-positive cognitively unimpaired individuals with preclinical AD, and Aβ-positive individuals with symptomatic disease. The aMTL_tau_-AT connections that were used in these group comparisons are shown to the left of the bar plot; the pMTL_tau_-PM connections are shown to its right. The AT network’s nodes and edges are in purple; those of the PM network are in green. Tau-based MTL ROIs are marked with a yellow asterisk. Abbreviations: AT = anterior-temporal; PM = posterior-medial; MTL = medial temporal lobe; aMTL_tau_ = tau-based anterior MTL ROI; pMTL_tau_ = tau-based posterior MTL ROI; CU = cognitively unimpaired; CI = cognitively impaired.

In summary, intra-network FC of both network systems displayed a progressive decline with age in Aβ-negative CU adults. The preclinical stage of AD, on the other hand, was characterized by increased functional synchronicity within the AT system, driven mainly by the aMTL_tau_-AT one-hop connections. Lastly, our results revealed that Aβ-positive individuals in the early stages of symptomatic AD displayed hypoconnectivity within the PM network.

## DISCUSSION

We conducted a detailed investigation of MTL FC changes in normal aging and preclinical and symptomatic AD. We paid particular attention to the anterior-posterior axis of the MTL to determine whether the anterior or posterior MTL segments were more vulnerable to age-or AD-related network dysfunction. Our most intriguing finding is the inverse ‘U-shaped’ FC pattern in the anterior MTL from normal aging to preclinical AD to symptomatic AD. The preclinical stage of AD was characterized by excessive FC centered around areas of the anterior MTL that are prone to early tauopathy. In patients with symptomatic disease, hyperconnectivity was no longer present. Instead, we observed reduced connectivity within the PM system and between various segments of the hippocampus. In contrast to the hyperconnectivity seen in preclinical AD, advanced age was associated with reduced intra-network FC in both the AT and PM systems. Within the MTL, we observed greater vulnerability of the posterior MTL, especially the PHC connections, to age-associated connectivity decline. Together, our results indicate that MTL network dysfunction in AD is not simply an accelerated form of normal aging.

### Hyperconnectivity in the Anterior Medial Temporal Lobe in Preclinical AD

In our earlier work, we reported increased FC between the ERC and other regions of the MTL in amnesic MCI.^43^ Later studies by others confirmed those findings.^22,45^ However, it was not obvious how early in the course of AD this effect first appears, and conflicting results have been published on the nature of the MTL network disruption at more advanced stages.^45,114^ Our results demonstrated that the anterior MTL hyperconnectivity is present in individuals with preclinical but not prodromal AD. Inverse ‘U-shaped’ FC patterns of this type have been previously identified in the default mode and salience networks.^115^ To our knowledge, only one other study reported elevated FC levels in the AT network among older individuals with memory complaints but not in patients with dementia due to AD.^114^ However, that study did not use PET or CSF biomarkers to screen for preclinical AD, and because of resolution limitations, it had limited anatomical specificity within the MTL.

The physiology underlying network hyperconnectivity in preclinical AD is unknown. Epidemiological studies indicate an elevated prevalence of subclinical seizures in both early-onset and late-onset AD,^116–118^ frequently originating in the temporal cortex and associated with earlier onset of cognitive symptoms.^119^ Even though we did not observe any differences in cognitive capacity between normal agers and CU participants with preclinical AD, it is plausible that increased anterior MTL connectivity in this group represents circuit dysfunction as opposed to compensatory processes. For instance, Bakker *et al*.^120^ showed that administering a low dose of the anti-epileptic drug levetiracetam to patients with amnesic MCI can result in memory improvement.

To our best knowledge, only a single human imaging study by Berron *et al*.^22^ examined the effects of preclinical and symptomatic AD on both the AT and PM networks. They reported decreased FC between the medial prefrontal cortex and the ERC/PRC region of the MTL in preclinical AD and decreased MTL-PM FC in Aβ+ individuals with amnesic MCI. Thus, it is plausible that functional abnormalities in preclinical AD are characterized not only by local and regional aMTL_tau_ hyperconnectivity but also by reduced connectivity between the anterior MTL and more distant regions of the neocortex. Furthermore, according to our current results, the MTL subregions most likely to show NFT accumulation are also the ones to display patterns of excessive connectivity in early stages of AD. Harrison *et al*.^121^ reached a similar conclusion for intra-HP connectivity, demonstrating a positive relationship between the intra-HP signal coherence (a measure of local connectivity) and tau burden in the ERC among CU older adults.

Animal and *in vitro* research has revealed that hyperactivity within the MTL enhances both amyloid and tau pathology in the HP and ERC,^48,53-55,122,123^ while the earliest stages of neuronal dysfunction in AD mice are characterized by Aβ-induced hyperactivity with hypoactivation appearing in later stages of the disease.^124^ Using mice models of AD, Angulo *et al*.^49^ studied the effects of Aβ and tau on ERC neurons specifically. Consistent with findings from non-ERC MTL subregions, Aβ – in the absence of tau – induced hyperexcitability in the ERC circuits; however, co-expression of both Aβ and tau moderated the excitatory effects of Aβ on the ERC.^49^ This is broadly consistent with our findings of increased connectivity in the anterior MTL only during the preclinical stage of AD when tau levels are expected to be low. Alternatively, by symptomatic stages of the disease, most individuals are expected to have a more significant tau burden both within the MTL and beyond.^12,16,55,125-128^

In this context, our results support the hypothesis that the earliest pathological processes in AD result in MTL network dysfunction centered around the anterior MTL subregions, possibly caused by Aβ-induced hyperexcitability. Such elevated activity levels likely accelerate NFT formation and spread, leading to synapse loss and network breakdown over time.

### Intra-MTL Functional Connectivity Changes in Normal Aging

We showed that posterior MTL subregions, and especially the PHC, become increasingly decoupled from the rest of the MTL with age. On standardized psychological assessments, older adults attain lower scores on tests of recollection and relational/associative memory, while familiarity-based item recognition is largely unaffected by age.^67–71,129^ Declining performance on tests of associative/relational memory indicates posterior, as opposed to anterior, MTL dysfunction in older adults.^6,7,130^ Consistent with the evidence from cognitive work, Panitz *et al*.^131^ reported reduced intra-HP connectivity in older participants, but only in the middle-posterior HP segments. Similarly, some structural MRI studies reported greater vulnerability of the posterior MTL subregions to atrophy in normal aging.^18,132–134^

In a task-based study of intra-MTL FC, Stark *et al*.^135^ compared PRC-HP, ERC-HP, and PHC-HP connectivity profiles of CU young (< 40 years) and CU old (≥ 70 years) adults. The authors reported reduced aHP-PHC connectivity among the elderly and a smaller but also significant reduction of the PRC-HP FC. However, group differences were not found in the more anterior ERC-HP connections. In the rs-fMRI literature, the effects of age on HP- and PHC-associated FC have been examined by Damoiseaux *et al*.,^136^ who reported negative age relationships for the anterior-posterior and interhemispheric intra-HP connections. By focusing on spontaneous network dynamics at rest and by studying all memory-related MTL subregions, not just the HP and PHC, we advanced the understanding about the effects of normal aging on intra-MTL FC.

### Effects of Age on Functional Connectivity Properties of the AT and PM Networks

This is the first FC study to document age-related alterations in the AT and PM intra- and inter-network connectivity for the entire adult lifespan. Initial imaging evidence for altered network dynamics in old age was demonstrated in task-based fMRI and PET experiments, which showed an over-recruitment of frontal and parietal association cortices in older cohorts relative to younger cohorts across a wide variety of cognitive tasks.^137–147^ Studies of spontaneous BOLD signal fluctuations also found a negative relationship between age and network specialization across standard cerebral networks, which often manifests as increased inter-network connectivity in older participants.^66,148–154^ Contrary to these observations, we did not detect any positive relationships between age and AT-PM inter-network connectivity. Instead, our results indicate reduced AT-PM interactions in older adults. This lack of agreement with earlier literature might be a consequence of our exclusive focus on the AT-PM connections. It is still plausible that the AT and PM networks undergo age-related despecialization, just not in their interactions with each other. Alternatively, MTL-affiliated network systems might be more resilient to the age-associated loss of functional specialization than other brain systems. Contrary to the inter-network results, our present findings on the AT and PM intra-network connectivity are broadly consistent with the aging literature from other groups, which also reported reduced intra-network interactions in cognitively normal older adults.^60,63–66,155,156^

### Limitations and Future Work

A few caveats merit discussion. First, as is the case with most fMRI studies of the MTL, the anterior MTL subregions in our study were more vulnerable to susceptibility artifacts from the skull base than their posterior counterparts. However, because both raw and preprocessed temporal signal-to-noise ratio (tSNR) profiles for each ASHS-T1 ROI were similar across groups (Suppl. Table 1), it is unlikely that susceptibility effects biased our main findings. Second, because of acquisition constraints, we assumed a common network architecture for all participants in a given statistical comparison. Despite high levels of structural consistency, inter-individual variability in network architecture exists,^157–159^ and exploring individual differences in the AT and PM network profiles at different stages of AD progression is a valuable avenue for future research.

Head motion has been shown to modulate FC in multiple brain networks.^160–162^ As in other studies involving older adults,^64,163^ head movement in our current study was correlated with age. Our preprocessing pipeline was designed with this issue in mind. Furthermore, among the three older groups [i.e., CU Aβ-, CU Aβ+, CI Aβ+] the amount of head motion was similar (Suppl. Table 2). Consequently, it is unlikely that our main findings were driven by head movement artifacts.

Conceptually, the present study focused only on the anterior-posterior axis of the MTL; however, work by others demonstrated functional specialization and differential AD pathology within the medial-to-lateral axis as well.^10,20,36,135,164–167^ Future studies employing high-field high-resolution fMRI acquisitions will be able to study the effects of AD progression on both axes simultaneously.

Although we infer opposite effects of Aβ and tau NFT on anterior MTL connectivity from our results, the proposed model needs further validation on participants who underwent both amyloid-PET and tau-PET scans. Lastly, the cross-sectional nature of our design – and all limitations that accompany cross-sectional studies of aging and AD – need to be acknowledged.

## CONCLUSION

Our results suggest that MTL-AT connectivity follows an inverse ‘U-shaped’ pattern in AD. The earliest phase of AD is characterized by elevated MTL-AT connectivity. This effect originates from the MTL regions that are most susceptible to early tauopathy. As the disease progresses, MTL-AT connectivity declines and hypoconnectivity appears in the PM network. In normal aging, both the AT and PM networks displayed intra-network connectivity decline; however, the deleterious effects of age were more pronounced in the posterior MTL subregions. Thus, MTL functional connectivity is a dynamic process in normal aging and in AD.

## Abbreviations

AD: Alzheimer’s disease
AT: anterior-temporal
BOLD: blood oxygen level-dependent
CI: cognitively impaired
CU: cognitively unimpaired
ERC: entorhinal cortex
FC: functional connectivity
fMRI: functional
MRI; HP: hippocampus
MTL: medial temporal lobe
PM: posterior-medial
PHC: parahippocampal cortex
PRC: perirhinal cortex

