## Supplemental materials for "Aging and Alzheimer’s Disease Have Dissociable Effects on Medial Temporal Lobe Connectivity"

### SUPPLEMENTARY METHODS

#### Anatomical Preprocessing in *fMRIPrep*

First, each participant's T1-weighted (T1w) image was corrected for intensity non-uniformity using the N4 algorithm (Tustison et al., 2010). Each T1w image was then skull-stripped with a Nipype (Gorgolewski et al., 2011) implementation of the ANTs brain extraction workflow, using OASIS30ANTs as the target template. Brain-extracted T1w scans were subsequently registered to the brain-extracted 1-mm MNI152NLin6Asym template using SyN diffeomorphic registration (ANTs 2.2.0; Avants & Gee, 2004; Avants et al., 2008), and brain surfaces were reconstructed using FreeSurfer 6.0.1 (Dale et al., 1999; http://surfer.nmr.mgh.harvard.edu). Subject-level CSF, WM, and gray matter (GM) segmentations were performed on brain-extracted T1-weighted scans using FSL *fast* (FSL 5.0.9; Zhang et al., 2001).

#### Amyloid PET Analysis

Amyloid PET data were processed with in-house software. First, attenuation-corrected dynamic image frames were motion-corrected using *mcflirt* rigid-body registration (FSL 5.0.9; Jenkinson et al., 2002; Smith et al., 2004). The resulting motion-corrected PET frames were averaged and aligned with participants' T1-weighted structural MRI scans using ANTs rigid-body registration with a mutual information metric (Avants & Gee, 2004; Avants et al., 2008). Each anatomical MRI was segmented into cortical, subcortical, and cerebellar ROIs using a multi-atlas segmentation method (Asman & Landman, 2013; Wang et al., 2013). Mean tracer uptake in the cerebellar gray and white matter was computed and used as a reference to generate a standardized uptake value ratio (SUVR) map for the entire brain. A composite ROI consisting of the middle frontal, anterior cingulate, posterior cingulate, inferior parietal, precuneus, supramarginal, middle temporal, and superior temporal cortical regions was used to compute a global SUVR for amyloid scans (Landau et al., 2013).

ROI-based Representation of the Extended MTL Network

Extra-MTL ROIs with positive functional connectivity (FC) to the MTL were selected from the 400-region 17-Network Schaefer et al. (2018) parcellation. Cortical regions with connectivity to the MTL were identified in normal agers only [i.e., CU young, middle-aged, and Aβ− older participants]. To test for the presence of FC to the MTL, we performed 4 sets (one per each MTL ROI: left anterior, right anterior, left posterior, right posterior) of one-sample positive-sided *t*-tests [FDR-corrected, $q < 0.05$] on Fisher-transformed subject-level Pearson correlation coefficients, representing that segment's FC to each of the 393 non-MTL Schaeffer ROIs. Seven Schaeffer ROIs were excluded because of substantial (>15%) spatial overlap with our anterior or posterior tau-based MTL seeds. To ensure that we did not miss any major cortical regions with FC to the MTL, we also performed seed-to-voxel network identification. Here, one-sample permutation tests (5,000 permutations) for positive connectivity to the bilateral anterior and posterior tau-based MTL ROIs were performed on Fisher-transformed subject-level voxelwise seed-to-voxel connectivity maps (Conn 20.b; Whitfield-Gabrieli & Nieto-Castanon, 2012). The Threshold-Free Cluster Enhancement (TFCE) method with the FDR ($q < .05$) correction for multiple hypothesis testing was used in these voxelwise tests (Benjamini & Hochberg, 1995; Smith & Nichols, 2009). We considered a given Schaeffer ROI as a part of the broader MTL-associated functional system if it was functionally connected to at least one of the MTL ROIs in the ROI-to-ROI network identification or if more than 40% of that ROI's voxels corresponded to a statistically significant cluster in the voxelwise network identification method. In total, we identified 221 Schaeffer ROIs with positive functional connectivity to the MTL. Together with 4 seed regions, these 221 Schaeffer ROIs (225 ROIs in total) were used in all subsequent analyses of the MTL network function (Fig. 2a in the main text).

**SUPPLEMENTARY FIGURES**

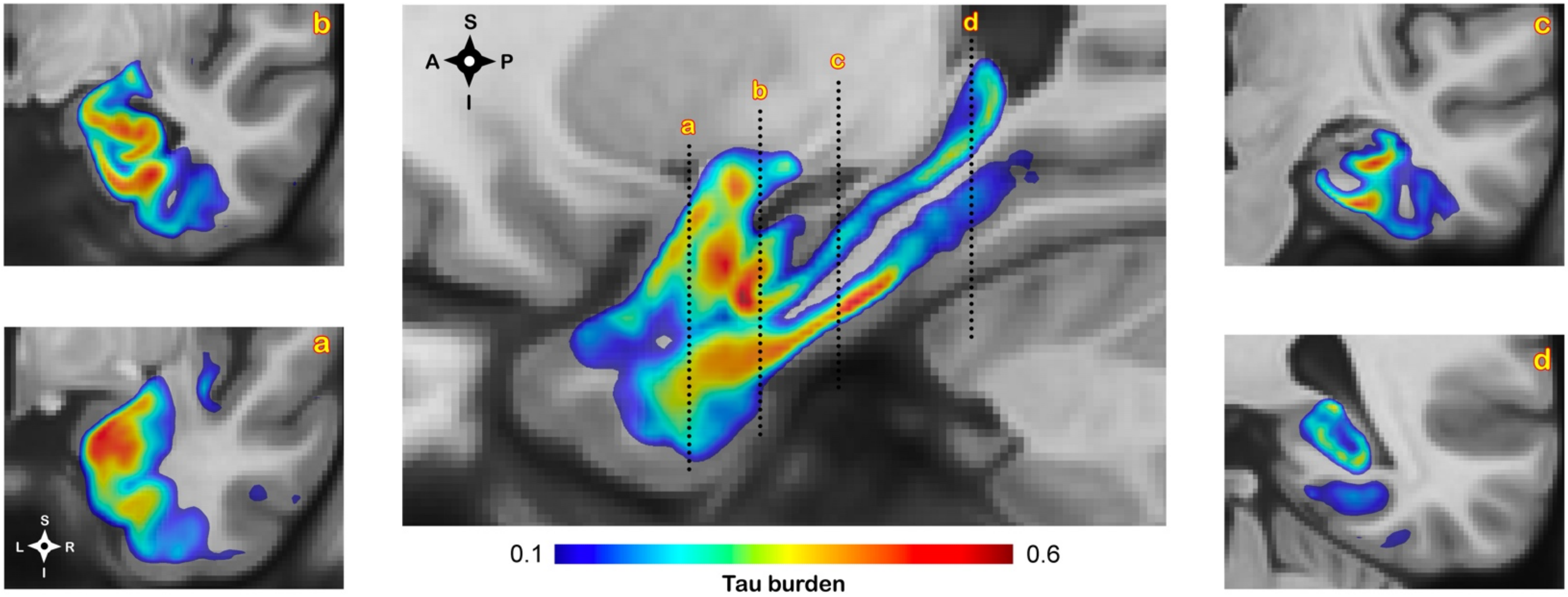


**Suppl. Figure 1**. Coronal and sagittal planes depict the overall Medial Temporal Lobe (MTL) neurofibrillary tangle burden, derived from a serial histological examination of 15 MTL specimens (for detailed methodology see, Yushkevich et al., 2021). These tau maps were used to create tau-based ROIs for MTL-AT and MTL-PM connectivity analyses.

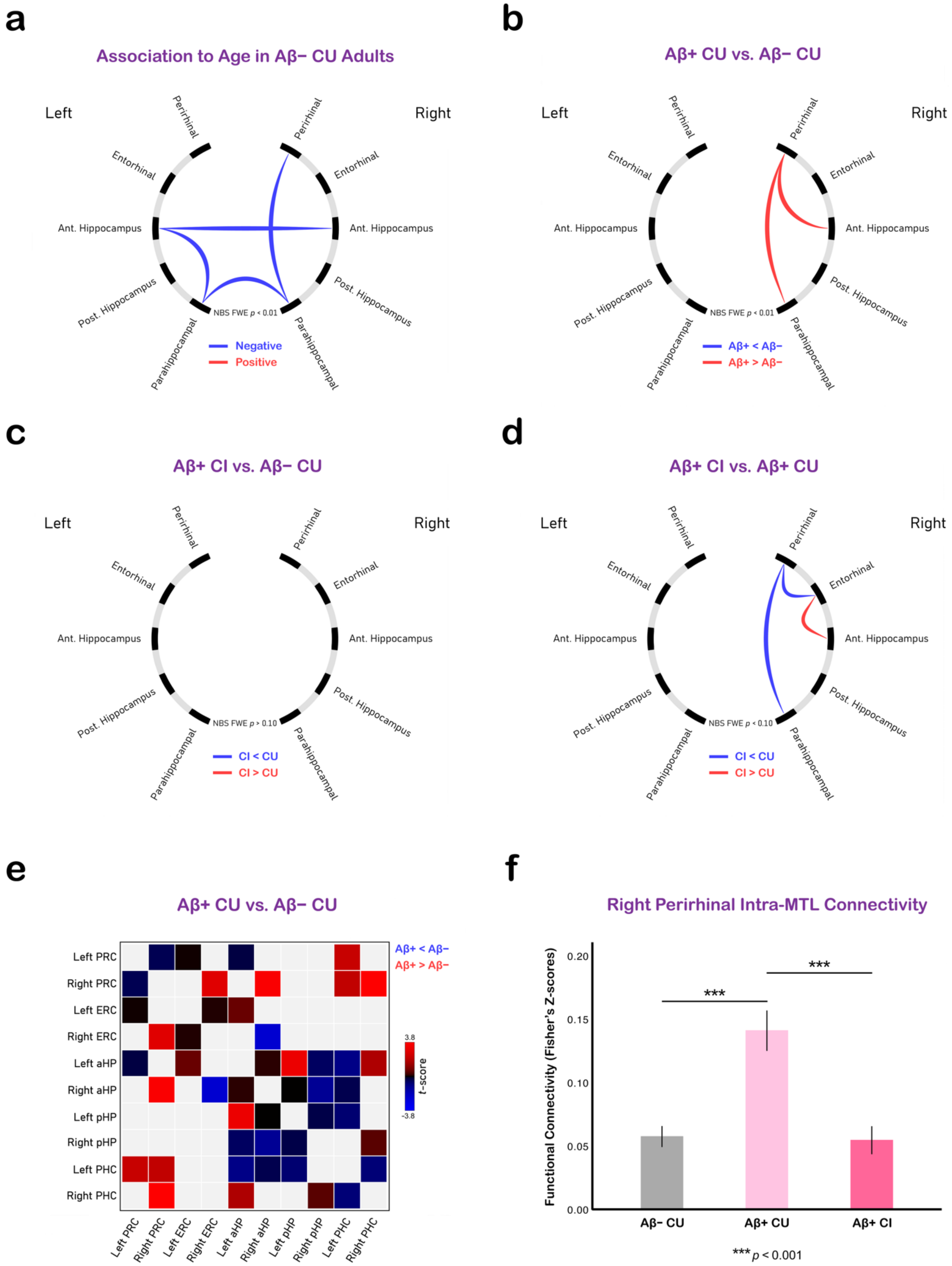


**Suppl. Figure 2**. This figure is a companion to Fig. 3 from the main text. Instead of canonical Pearson correlations, intra-MTL functional interactions were quantified using partial correlation coefficients, controlling for time courses from all other ASHS-T1 ROIs. Only those connections that represented *direct* intra-MTL functional interactions (see Fig. 2b in the main text) were analyzed. Connectograms depict the effects of **(a)** age and **(b-d)** AD progression on *direct* intra-MTL functional connectivity. **(e)** Matrix-form representation of *direct* intra-MTL connectivity differences between Aβ-positive individuals with preclinical AD and Aβ-negative age-matched controls. **(f)** Average strength of *direct* connections from the right PRC to other ASHS-T1 ROIs, in Aβ-negative normal agers, Aβ-positive cognitively normal individuals with preclinical AD, and Aβ-positive individuals with symptomatic disease. Abbreviations: PRC = Perirhinal Cortex; ERC = Entorhinal Cortex; PHC = Parahippocampal Cortex; aHP = Anterior Hippocampus; pHP = Posterior Hippocampus; CU = cognitively unimpaired; CI, cognitively impaired.

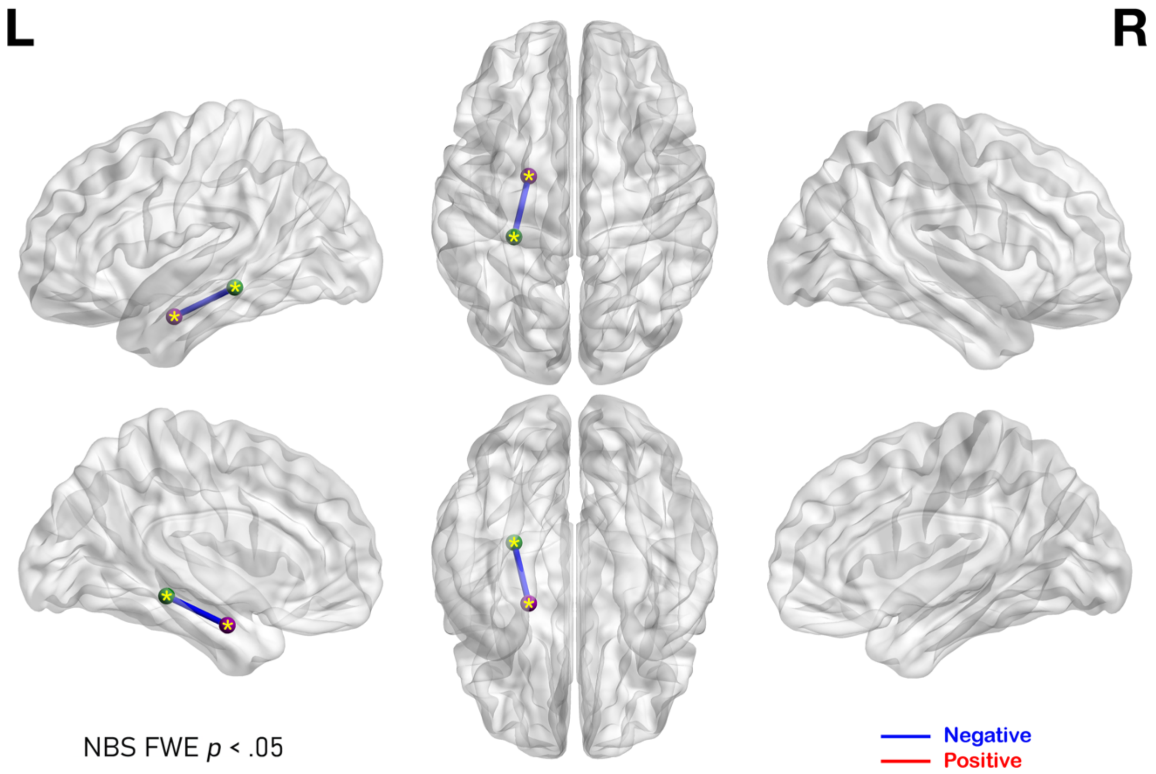


**Suppl. Figure 3**. The effect of normal aging on inter-module AT-PM connectivity. A single connection, linking the anterior and posterior tau-based MTL ROIs in the left hemisphere with each other, showed connectivity decline in amyloid-negative cognitively unimpaired agers.

**SUPPLEMENTARY TABLES**

**Suppl. Table 1.** MTL voxelwise temporal Signal-to-Noise Ratio (tSNR) for raw and preprocessed fMRI datasets, separated by group.

| | PRC | | ERC | | PHC | | Anterior HC | | Posterior HC | | Anterior Tau | | Posterior Tau | |
|---|---|---|---|---|---|---|---|---|---|---|---|---|---|---|
| | Left | Right | Left | Right | Left | Right | Left | Right | Left | Right | Left | Right | Left | Right |
| *Raw fMRI Data* | | | | | | | | | | | | | | |
| Young and Middle-Aged | 13.63 (2.62) | 13.44 (2.34) | 9.55 (2.20) | 10.15 (2.16) | 21.90 (2.57) | 21.62 (2.31) | 16.97 (3.27) | 18.10 (3.18) | 21.74 (2.33) | 21.82 (2.20) | 14.27 (2.03) | 15.49 (2.06) | 20.83 (2.74) | 20.60 (2.50) |
| Aβ- CU | 12.48 (2.49) | 11.91 (2.42) | 8.92 (2.08) | 9.29 (2.57) | 19.45 (3.13) | 18.86 (3.09) | 16.14 (2.65) | 16.81 (2.77) | 20.61 (2.22) | 20.35 (2.34) | 13.55 (2.09) | 14.41 (2.28) | 19.52 (2.55) | 19.12 (2.60) |
| Aβ+ CU | 12.70 (2.08) | 12.29 (2.34) | 7.92 (1.86) | 8.94 (2.44) | 19.78 (2.38) | 19.73 (2.52) | 15.69 (3.48) | 16.73 (3.61) | 20.77 (2.19) | 20.90 (2.18) | 13.14 (1.91) | 14.08 (2.51) | 19.83 (2.25) | 19.70 (2.01) |
| Aβ+ CI | 12.00 (2.10) | 11.56 (2.03) | 7.95 (1.54) | 8.90 (2.21) | 19.13 (2.08) | 18.61 (2.07) | 15.33 (2.57) | 15.99 (3.12) | 20.47 (1.90) | 20.36 (1.96) | 13.63 (1.69) | 14.43 (2.07) | 19.44 (2.10) | 19.33 (2.04) |
| *Preprocessed fMRI Data* | | | | | | | | | | | | | | |
| Young and Middle-Aged | 263.3 (51.5) | 260.5 (51.3) | 194.9 (40.1) | 202.4 (34.5) | 351.4 (47.3) | 344.0 (45.7) | 303.5 (58.6) | 317.4 (55.2) | 368.6 (41.4) | 373.2 (39.8) | 262.9 (46.8) | 280.2 (44.9) | 364.2 (51.0) | 364.5 (44.1) |
| Aβ- CU | 247.1 (47.3) | 236.6 (48.9) | 192.5 (32.4) | 198.7 (35.8) | 326.7 (38.9) | 319.6 (46.8) | 300.0 (42.2) | 309.0 (42.6) | 362.2 (39.2) | 357.2 (38.8) | 257.4 (38.0) | 268.8 (38.9) | 354.2 (45.5) | 347.3 (47.0) |
| Aβ+ CU | 244.8 (44.3) | 240.0 (51.4) | 176.9 (33.0) | 190.5 (39.6) | 322.1 (33.4) | 321.5 (30.8) | 296.8 (48.1) | 309.7 (53.2) | 362.3 (40.6) | 365.5 (32.7) | 253.4 (39.6) | 264.8 (49.2) | 354.8 (40.4) | 352.7 (34.6) |
| Aβ+ CI | 243.0 (43.4) | 232.8 (42.4) | 193.8 (28.1) | 198.4 (40.0) | 337.2 (42.4) | 326.9 (35.4) | 317.7 (47.7) | 320.9 (52.3) | 381.2 (46.9) | 384.7 (40.7) | 272.9 (38.0) | 280.2 (41.9) | 374.8 (51.7) | 373.3 (46.6) |

**Suppl. Table 2.** Displacement (FD) for raw and filtered realignment parameters, separated by group.

| | Young and Middle-Aged (mm) | Aβ- CU (mm) | Aβ+ CU (mm) | Aβ+ CI (mm) |
|---|---|---|---|---|
| Raw Mean FD (SD) | 0.1588 (0.0700) | 0.2627 (0.1236) | 0.2303 (0.0814) | 0.2158 (0.0974) |
| Raw Max FD (SD) | 0.6209 (0.5888) | 1.1046 (1.0069) | 1.0697 (0.7293) | 1.2868 (1.0637) |
| Filtered Mean FD (SD) | 0.0213 (0.0118) | 0.0403 (0.0193) | 0.0420 (0.0173) | 0.0401 (0.0263) |
| Filtered Max FD (SD) | 0.1698 (0.1354) | 0.2273 (0.1900) | 0.2633 (0.1737) | 0.2915 (0.2155) |
